# Performance of a multi-target, multi-cancer early detection (MCED) blood test in a prospectively collected cohort

**DOI:** 10.1101/2025.08.24.25334244

**Authors:** Vladimir Gainullin, Mael Manesse, Fanglei Zhuang, Melissa Gray, Hee Jung Hwang, Gustavo C. Cerqueira, Christopher L. Nobles, Madhav Kumar, Kevin Arvai, Xi Chen, Christopher C. Tyson, Chen Ji, Jin Bae, Violeta Beleva Guthrie, Jaspreet Kaur, Viatcheslav E. Katerov, Larson Hogstrom, Leonardo Hagmann, Philip J. Uren, Jorge Garces, Hatim T. Allawi, Vuna Fa, Abigail McElhinny, Gerard A. Silvestri, David Chesla, Robert W. Given, Tomasz M. Beer, Frank Diehl

**Affiliations:** Exact Sciences Corporation, Madison, WI; Exact Sciences Corporation, La Jolla, CA; Medical University of South Carolina, Charleston, SC; Corewell Health, Grand Rapids, MI; Urology of Virginia, Virginia Beach, VA

**Keywords:** Multi-cancer early detection, cell-free DNA (cfDNA), methylation, proteins, mutations, reflex assay, machine learning

## Abstract

**Background:** Multi-target blood tests have the potential to detect a broad range of cancer types and stages. We previously trained and independently assessed the performance of four biomarker classes for cancer detection in a retrospectively assembled case-control feasibility study. The primary aim of this study was to assess the performance of a methylation and protein (MP) classifier using samples from a large, multi-center, prospectively collected case-control study. As part of the ongoing assay development process, we also performed an exploratory analysis investigating the addition of a somatic mutation reflex approach (MP-reflex) on samples with scores below the threshold for a positive MP call but above a clear negative MP call.

**Methods:** This study included 6,352 samples (1,438 cancer and 4,914 non-cancer) representing all stages among 21 tumor organ types divided approximately equally between a training and a test set. The sensitivity of the MP classifier was determined at a high specificity. The MP-reflex approach was investigated on MP samples at the same specificity threshold and detectable MP signals above pre-specified lower thresholds.

**Results:** At 98.5% specificity, the observed overall sensitivity for MP biomarkers was 50.9%, with sensitivities of 15.4%, 26.1%, 38.0%, 67.8%, 85.5%, and 37.5% for stages I, I and II combined, II, III, IV, and unknown, respectively. When breast and prostate cancers were excluded from the analysis, overall test sensitivity was 56.8%, with sensitivities of 17.2%, 30.7%, 48.6%, 73.5%, 86.5%, and 40.0% for stages I, I and II combined, II, III, IV, and unknown, respectively.

**Conclusion:** Methylation and protein biomarkers detected all cancer organ types incorporated in the analysis, including those without standard of care (SoC) cancer screening options and aggressive cancers with poor 5-year survival. These results demonstrate the potential clinical validity of multi-biomarker testing for the detection of several cancer types.

## Introduction

Cancer is the second leading cause of death in the U.S, with more than two million new cancer cases and over 618,000 cancer deaths projected to occur in 2025.^1^ The United States Preventive Services Taskforce (USPSTF) recommends routine cancer screening for breast, colorectal, and cervical cancer in average risk individuals, and for lung cancer in individuals with elevated risk due to a pre-specified threshold exposure to tobacco.^2–5^ Standard of care (SoC) cancer screening programs have improved mortality rates for these four cancers; however, two-thirds of annual cancer deaths are the result of cancers without endorsed screening.^1, 6^

Multi-cancer early detection (MCED) testing has the potential for earlier detection of multiple cancers simultaneously, including lower prevalence cancers and aggressive cancers (i.e. pancreatic, esophageal, liver, stomach, and ovarian) that are often detected at later stages.^1, 6^ Advances in molecular and computational technologies have enabled the sensitive and specific detection of tumor-derived biomarkers in the blood. Most MCED assays currently in development utilize single biomarker classes, such as cell-free DNA (cfDNA) hyper-methylation,^7–14^ mutations,^15^ fragmentation patterns,^16–20^ or proteins^21^ to detect cancers.

However, the utilization of single biomarker types may not account for the complexity and heterogeneity of cancer, particularly when tasked with detecting multiple cancers by a single test.^22^ A combined, multi-target approach provides a more comprehensive assessment of shared cancer-associated biomarkers present in the blood, with the goal of improved sensitivity for early cancer detection.^23^ To that end, MCED tests incorporating multi-biomarker classes are also in development, utilizing DNA fragmentation and methylation with or without protein markers, or cfDNA mutation, aneuploidy, and methylation biomarkers, together with cancer-associated protein quantification.^24–29^

CancerSEEK was a multi-target MCED assay developed in a case-control setting and examined in a large prospective study and served as the foundation for subsequent test development. CancerSEEK (mutations and select proteins) was utilized in the <u>D</u>etecting cancers <u>E</u>arlier <u>T</u>hrough <u>E</u>lective mutation-based blood <u>C</u>ollection and <u>T</u>esting (DETECT-A) study; the first prospective interventional clinical study to evaluate an MCED blood test. In DETECT-A, CancerSEEK detected 19% of stage I cancers, 15% of stage II cancers and 30% of stage III cancers at a specificity of 99%.^27^ Using another initial version of CancerSEEK (mutations, proteins, and a machine learning classifier), Cohen et al., reported a sensitivity of 70% and a specificity of over 99% for eight types of cancer (ovary, liver, stomach, pancreas, esophagus, colorectal, lung, or breast) in a case-control feasibility study of 1005 patients.^24^

Douville *et al.* hypothesized that the test performance observed in the Cohen study could be improved by the addition of aneuploidy to the biomarker panel.^30^ While maintaining a specificity of 99%, the combination of aneuploidy, cfDNA mutations, proteins, and a machine learning classifier detected seven cancer types (liver, ovary, pancreas, esophagus, stomach, colorectal, and lung); the sensitivity ranged from 77 to 97% and 38% for breast cancer. Using retrospectively assembled case-control feasibility study data, our group has also assessed MCED test performance utilizing DNA methylation and protein combinations,^31, 32^ as well as 3-biomarker classes (methylation, aneuploidy, and proteins) and 4-biomarker classes (methylation, mutations, aneuploidy, and proteins) with a machine learning classifier.^25, 26^ The aim of this study was to assess the performance of a two-biomarker (methylation and protein, MP) MCED assay using samples from a large, multi-center, prospectively collected case-control study. Aneuploidy was tested, but we did not include a performance assessment in this study (unpublished data). We also assessed the performance of a somatic mutation reflex approach in samples with initially negative MP-results (MP-reflex) to test the feasibility of an MP-reflex assay in a high-throughput, reduced cost setting. We hypothesized that MP-reflex might be an efficient strategy to add a third biomarker class and enhance sensitivity for the detection of early-stage cancer.

## Methods

### Study Participants and Sample Collections

Ascertaining Serial Cancer patients to Enable New Diagnostic 2 (ASCEND-2), a multi-center, prospectively collected case-control study of clinically characterized participants, was the main source of the samples for this classifier development study. One hundred ninety sites within the United States (35 states) and 15 sites within Europe were engaged for subject enrollment.

ASCEND-2 study included male and female subjects ≥50 years old with known cancer, suspicion of cancer, and controls without suspicion of cancer. Cancer cases had untreated primary malignancies confirmed through pathology reports and/or clinical/radiographic data. Subjects were excluded from the study if they had any previous cancer diagnosis within the past 5 years (with the exceptions of basal cell or squamous cell skin cancers), or cancer recurrence within the study timeframe, or a concurrent diagnosis of multiple primary cancers. While free from suspicion of cancer at the time of enrollment, controls were not required to have completed all recommended cancer screening tests and were not followed contributing a blood sample to confirm the absence of cancer. All collection sites had Institutional Review Board approval, and all eligible subjects provided written informed consent and were assessed for study participation eligibility. Over 25,900 participants were enrolled in ASCEND-2, which included over 5,900 known cancer cases. A subset of these samples was selected to develop two different MCED cancer classifiers. Samples were selected based on plasma volume availability, clinical eligibility criteria, availability of validated clinical data prior to laboratory testing initiation, and demographic matching requirements between the training and test set cohorts. The number of cancers per organ site was targeted to represent cancers with high, common, and rare incidence compared to adjusted National Cancer Institute Surveillance, Epidemiology, and End Results (SEER) cancer incidence (accessed March 2024).^6^ The number of cancers per stage was targeted to be equal and not represent SEER to allow for data generation across all stages. In order to meet these sample selection targets the ASCEND-2 enrollment was supplemented with a set of 187 samples from a commercial vendor (Audubon Bioscience, Houston TX), and 12 cancer samples from an Exact Sciences study, Specimen Collection Study to Evaluate Biomarkers (2021-05). Blood was collected in LBgard^®^ tubes for plasma and buffy coat extraction.

### Methylated DNA Marker (MDM) Analysis

MDM candidates were identified from a genome-wide discovery approach using reduced representation bisulfite sequencing of DNA extracted from frozen primary cancer tissues and control tissues, in addition to normal buffy coat control samples. Candidate regions were selected where there was differential hypermethylation between the cases and controls. MDM candidates were then confirmed biologically on independent tissues before being tested on archived plasma. The results of these marker selection and elimination studies were designed primarily to remove non-discriminating biomarkers. Only the top performing MDMs across the many different cancer studies were chosen for further evaluation as candidate methylation markers for blood-based detection of cancer.^31, 33^

cfDNA was extracted from 6 mL plasma followed by bisulfite conversion on a Hamilton Microlab STARlet (Hamilton Robotics, Reno, NV) using reagents manufactured by Exact Sciences Corporation (Madison, WI). Bisulfite converted cfDNA was then analyzed using Target Enrichment Long-probe Quantitative Amplified Signal (TELQAS^TM^) chemistry.^31, 33^ For the initial target enrichment, converted cfDNA was transferred to a 96-well polymerase chain reaction (PCR) plate and subjected to a multiplex PCR to amplify the MDMs and a reference methylation marker. The PCR products were diluted 10-fold and evaluated in triplex or biplex assays using Long-probe Quantitative Amplified Signal (LQAS^TM^) on a Quantstudio 5 Dx Real-Time PCR Instrument (Life Technologies Corporation, Grand Island, NY). TELQAS combines amplification using real-time PCR and allele-specific detection of methylated target DNA through an invasive cleavage assay.

### Protein Analysis

Plasma protein concentrations were measured using commercially available assays for select proteins on a Roche cobas e801 instrument (Roche Diagnostics, Indianapolis, IN) per manufacturer recommendations.^33^ The cobas e801 analyzer uses electrochemiluminescence (ECL)-based immunoassays to measure protein concentrations.

### Mutation Analysis

Select mutations (i.e. somatic variants associated with cancer) were assessed by extracting cfDNA from ∼10 mL of plasma followed by sequencing using a modified version of a technology described by Cohen et al. 2021.^34^ Genomic DNA (gDNA), extracted from the matched buffy coat, was used for sequencing regions that were positive for mutations in cfDNA to verify whether the detected mutations were attributable to clonal hematopoiesis of indeterminate potential (CHIP). Only mutations detected in plasma but absent in the matched buffy coat samples were considered tumor derived variants. Samples were subjected to quality control (QC) based on sample-level, run-level, and control-level metrics. Sample counts and QC failure modes were recorded. The MP and MP-reflex testing workflow is shown in Figure 2.

### Machine learning model development MP classifier development

Separate plasma aliquots were tested with the protein and methylation assays as illustrated in Figure 2. Prior to classifier training, samples were partitioned into training (cross-validation (CV), mini-hold-out) and test (hold-out test) sets (Figure 1). Only samples that passed pre-specified methylation and protein assay quality control criteria were included in the training process. The training set was used to select model architecture, feature engineering, and transformation approaches as well as to perform hyperparameter tuning, and calibration of specificity thresholds. MP classifier threshold calibration was performed on the CV set, with the aim of achieving greater than or equal to 99.0% specificity while maintaining high sensitivity within a mini-hold-out set (223 cancers; 985 non-cancers).

**Figure 1.**
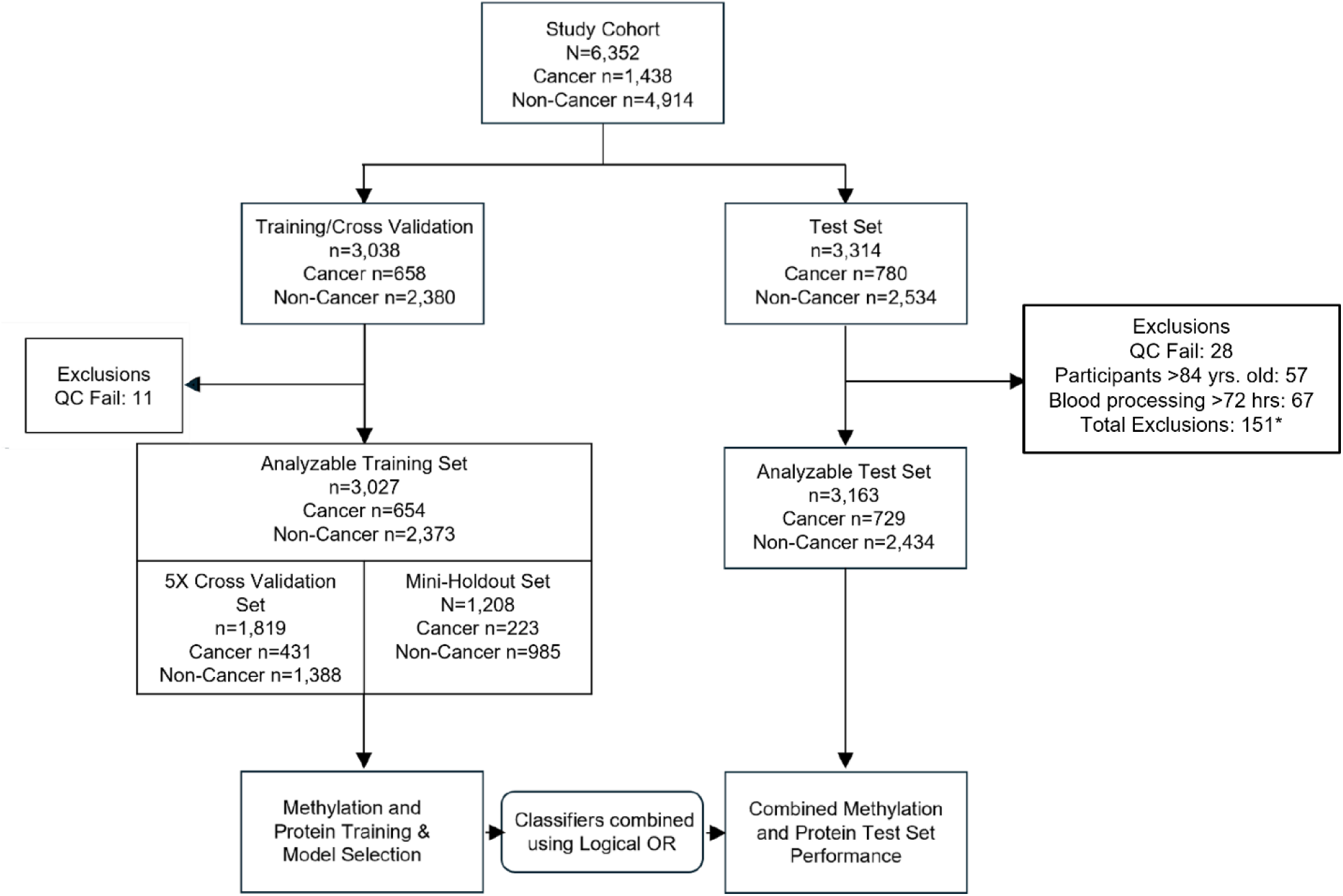
MP classifier development and test cohorts. MP, methylation-protein. *One sample was excluded for both age and blood processing reasons.

**Figure 2.**
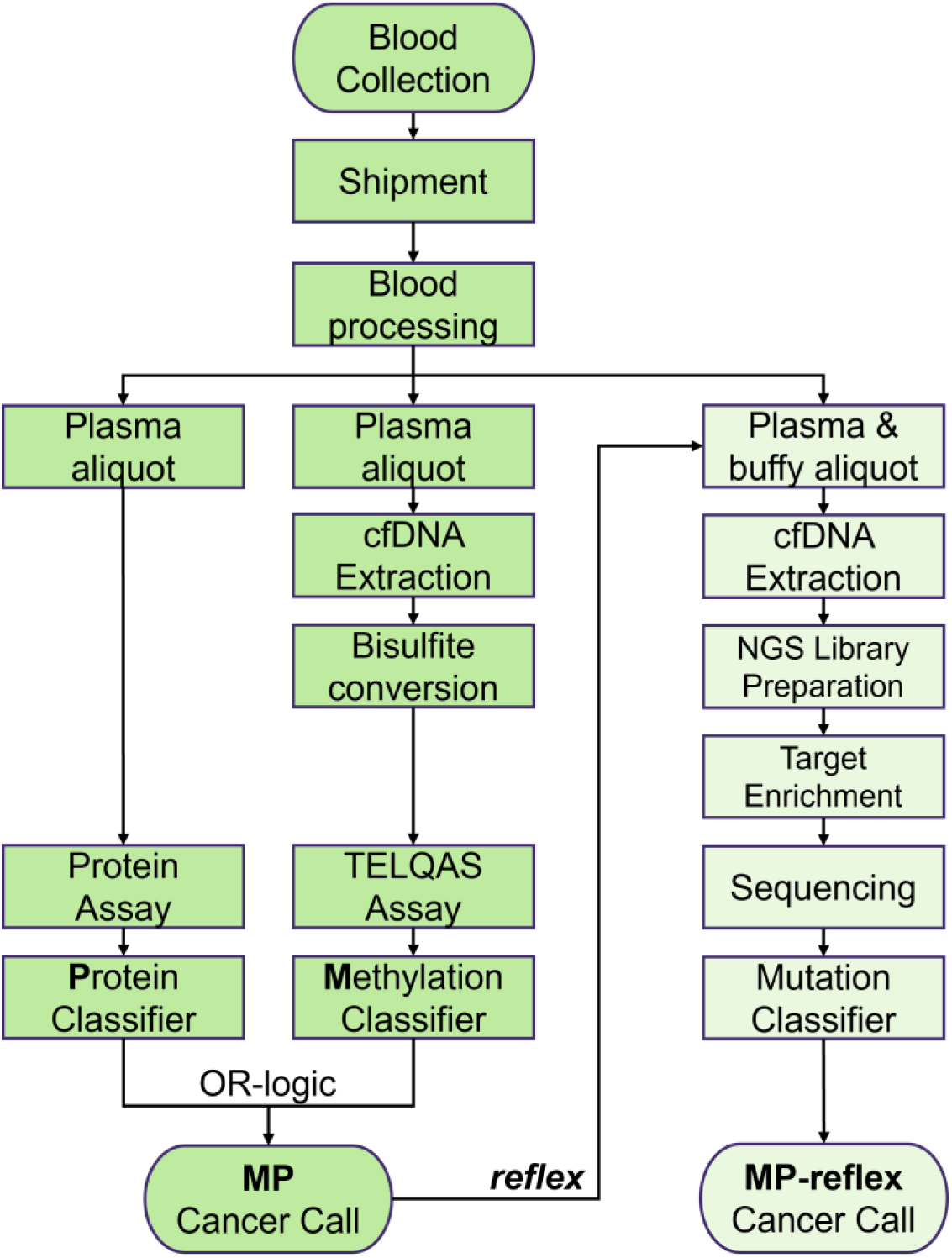
Testing workflow. LR, logistic regression; MP, methylation-protein.

Machine learning was used to develop and train two individual binary (cancer or non-cancer) classifiers for the methylation and protein panels, respectively (Figure 3a). The overall methodology was as follows: 5-fold stratified CV partitions were generated using age bin (half-decade), gender, ethnicity, race, cancer organ and stage as strata with the aim to produce proportional train/validate partitions across folds. These static partitions were then used to perform model selection across common linear and non-linear estimator architectures. Model bias estimation was evaluated in comparison between train/validation performance using sensitivity, specificity, total and partial Receiver Operating Characteristic Area Under the Curve (ROC AUC) results. Model architecture with the least bias and highest performance was further refined with hyper-parameter tuning and validation curve assessment. Robust model parameters and thresholds were then locked before final model performance assessment using the independent held-out test set.

**Figure 3.**
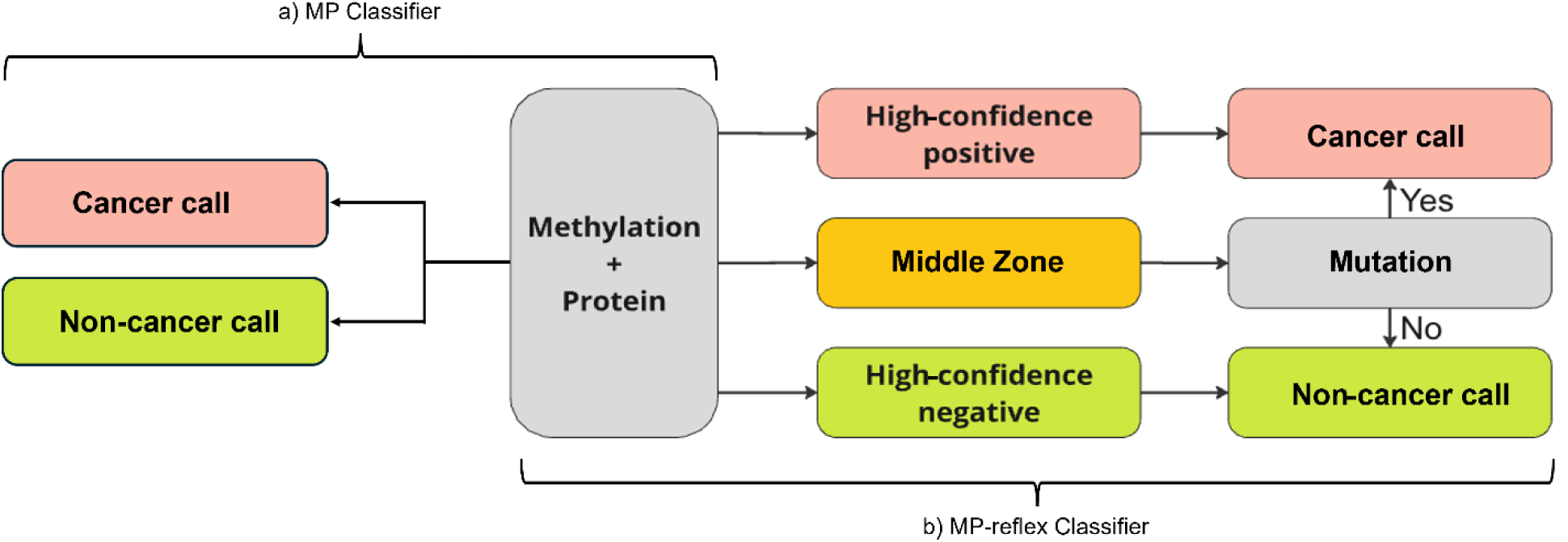
a) Methylation-Protein and b) Methylation-Protein-reflex classifier schema. MP, methylation-protein.

After classifier development was completed, the individual methylation and protein classifier results (cancer or non-cancer calls) were combined by an OR-logic overarching classifier to generate the final classification status (positive or negative). OR-logic was selected as the overarching MP classifier; other approaches (early and late fusion, weighted voting, and logistic regression) did not offer significant advantages. The result was deemed positive when either the methylation or protein assay provided a cancer-positive call. Hold-out test set samples were evaluated using this final overarching OR-logic classifier configuration (Figure 3a).

### MP-reflex classifier development

All samples were analyzed using the combined MP classifier described above. The MP-reflex classifier training included 20 organ sites and all stages of cancer. Mutation classifier thresholds were tuned using optimization in cross-validation within the training data set. Samples were tested following the mutation assay workflow using plasma as well as the corresponding buffy aliquots. The schematic of the different cancer classifiers that were developed and tested is shown in Figure 3.

The mutation classifier consisted of three main components: variant-level artifact score, CHIP, and sample-level aggregate mutation score that weighed evidence from variants passing artifact and CHIP filter. The three components were developed using an independent sample collection. The MP classifier was adapted to determine three outcomes (three-zone partitioning): high-confidence cancer positive (red), high-confidence cancer negative (green), and signal that fell between these two categories or zones (yellow; middle) (Figure 3b). Samples with high-confidence scores were classified as cancer or non-cancer calls, respectively. Samples with MP results that fell between high-confidence positive or high-confidence negative results (e.g., middle zone) underwent a follow up mutation assay evaluation. An OR logic model was chosen as the overarching MP classifier.

For MP-reflex classifier training, mutation samples that failed QC and were in the middle zone were considered negative (no variants) to avoid biasing the MP-reflex training results from multiple mutation configuration versions to MP. MP and MP-reflex thresholds were calculated using ROC curves in stratified cross-validation. Optimal thresholds across validation folds were averaged, locked before testing and used for final performance estimation. The resulting mutation classifier differentiated the outcome into a binary cancer or non-cancer call.

### Statistical analysis

Sensitivity, specificity and binary classification thresholds were calculated using scikit-learn metrics. 95% confidence intervals were calculated using binomial proportion Wilson score method (statsmodels).

## Results

### Participants and Sample Characteristics

A total of 6,352 samples were selected from the ASCEND-2 study as well as two other collections to develop and test a MP (Figure 1) as well as an MP-reflex cancer classifier (Figure 5). This overall set included 1,438 cancer participants across 21 organ sites and stages (anus (n=28), bladder and urinary (n=65), breast (n=165), cervix uteri (n=22), colon and rectum (n=161), esophagus (n=53), head and neck (squamous cell) (n=78), uterine (n=81), kidney (n=80), lung and bronchus (n=348), pancreatic (n=76), prostate (n=66), small intestine (n=12), liver and bile duct (n=50), stomach (n=58), ovary (n=37), thyroid (n=25), vulva (n=21), testis (n=2), and hematologic malignancies (non-Hodgkin lymphoma (NHL; n=9) and multiple myeloma; n=1) (Supplementary Materials Table 1). Non-cancer and cancer participants were similar for age, sex, and race/ethnicity distributions (Table 1). The study recruited participants that were broadly representative of the US population, with 81.7%, 13.0%, 12.9%, and 3.2%, identifying as White, Hispanic/Latino, Black/African American, and Asian, respectively.

**Table 1.** Participant demographics for classifier development.

| Variable |  | Non-Cancer<br>(4914) | Cancer<br>(1438) | Total (6352) |
| --- | --- | --- | --- | --- |
| Sex | Female | 2799 (57.0%) | 776 (54.0%) | 3575 (56.3%) |
|  | Male | 2115 (43.0%) | 662 (46.0%) | 2777 (43.7%) |
| Age (years) | Mean (SD) | 65.1 (8.1) | 66.9 (9.0) | 65.5 (8.3) |
|  | (Min, Max) | (50, 101) | (50, 92) | (50, 101) |
| Race | American Indian or<br>Alaska Native | 20 (0.4%) | 9 (0.6%) | 29 (0.5%) |
|  | Asian | 145 (3.0%) | 58 (4.0%) | 203 (3.2%) |
|  | Black or African<br>American | 698 (14.2%) | 119 (8.3%) | 817 (12.9%) |
|  | Mixed Race | 11 (0.2%) | 2 (0.1%) | 13 (0.2%) |
|  | Native Hawaiian or<br>Other Pacific<br>Islander | 5 (0.1%) | 1 (0.1%) | 6 (0.1%) |
|  | Unknown/Missing | 39 (0.8%) | 56 (3.9%) | 95 (1.5%) |
|  | White | 3996 (81.3%) | 1193 (83.0%) | 5189 (81.7%) |
| Ethnicity | Hispanic/Latino | 748 (15.2%) | 80 (5.6%) | 828 (13.0%) |
|  | Not Hispanic/Latino | 4135 (84.1%) | 1308 (91.0%) | 5443 (85.7%) |
|  | Unknown/Missing | 31 (0.6%) | 50 (3.5%) | 81 (1.3%) |

Specifically, based on the 2020 U.S. census rates and the FDA’s 2020 clinical trial participation rates,^35, 36^ the Black/African American population was slightly overrepresented in both the training and testing cohorts. The Hispanic/Latino ethnicity rate exceeded the clinical trial participation rate; whereas the representation rate for Asian participants was slightly lower than both the 2020 U.S. census and FDA 2020 clinical trial participation rates. Overall, the race/ethnicity participation rates aligned with expectations for a classifier development study.

Cancer stages and organ sites in the training and test sets collectively represent more than 80% of incident cancers observed in the general population.^1^ The number of cancers per organ site was targeted to represent cancers with high, common, and rare incidence compared to adjusted SEER cancer incidence (accessed March 2024),^6^ lung and bronchus cancers were over-represented (SEER: 15.8% vs. 24.2% for ASCEND-2), while breast (17.7% vs. 11.5%), prostate (17.5% vs. 4.6%), and hematological cancers were under-represented. The cancers used for the classifier development by cancer organ type as shown in Supplementary Table 1. Participant demographics for the analyzable MP training and test sets are shown in Supplementary Table 2. Overall, cancer stage distributions within organ sites were approximately equal for stages I-IV (27%, 21%, 26%, and 22% respectively) and ∼3% with unknown stage (Supplementary Figure 1).

### Methylation-Protein (MP) classifier training and testing

#### MP classifier interrogation

The generalizability of the MP classifier was evaluated by comparing the performance of the full training set (including the CV set and mini-hold-out set), the CV set alone, and the test set. The full training set (n=3,027) and 5-fold cross-validation set (n=1,819) achieved 98.9% (95% CI: 98.1-99.3%) and 98.8% (95% CI: 98.3-99.2%) specificity, respectively. The test set (n=3,163) achieved 98.5% specificity (95% CI: 97.9-98.9%), with a false positive rate of 1.5% (37/2432 non-cancer cases). No significant bias was observed between the full training set, CV set, and test set performance by stage (Supplementary Figure 2) or specific tumor organ site (Supplementary Figure 3).

### MP classifier test set performance

The test set included 780 cancers and 2,534 non-cancers that were all tested using the methylation and protein assays. A total of 28 (0.8%) samples failed pre-specified assay quality control criteria. Fifty-seven samples from participants over 84 years of age (outside of the intended use age range) were removed from the test set analysis. A 72-hour blood processing window requirement was also applied to the test set cohort that resulted in the exclusion of an additional 67 cases. MP classifier test set performance without age and blood processing exclusions is shown in Supplementary Table 5. The set without these exclusions demonstrated an overall sensitivity of 51.6% (95% CI: 48.0-55.1%) at a specificity of 98.4% (95% CI: 97.9-98.9%).

The remaining analyzable MP sample set was 729 cancers and 2,434 non-cancers. The MP classifier demonstrated an overall sensitivity of 50.9% (95% CI: 47.3-54.5%), and sensitivities of 15.4% (95% CI: 10.9-21.3%), 26.1% (95% CI: 21.7-31.0%), 38.0% (95% CI: 30.9-45.7%), 67.8% (95% CI: 60.6-74.2%), 85.5% (95% CI: 79.4-90.0%) and 37.5% (95% CI: 22.9-54.7%) for stages I, I and II combined, II, III, IV, and unknown, respectively for 21 organ types (Table 2). MP classifier performance was also assessed with breast and prostate cancers excluded, with an observed overall sensitivity of 56.8% (95% CI: 52.8-60.7%), and sensitivities of 17.2% (95% CI: 12.0-24.2%), 30.7% (95% CI: 25.4-36.6%), 48.6% (95% CI: 39.4-57.9%), 73.5% (95% CI: 66.0-79.9%), 86.5% (95% CI: 80.2-91.0%), and 40.0% (95% CI: 24.6-57.7%) for stages I, I and II combined, II, III, IV, and unknown, respectively. Overall sensitivity excluding cancer organ types with average-risk SoC screening (i.e. excluding breast, prostate, cervix uteri, colon and rectum) was 54.8% (95% CI: 50.4-59.2%) and 63.7% (95% CI: 58.0-69.0%) for the six most aggressive cancer organ types with the shortest 5-year survival rate^1^ (i.e. pancreas, esophagus, liver, lung and bronchus, stomach, and ovary). MP organ-specific performance ranged from 11.8% (95% CI: 5.5-23.4%) for prostate to 80.0% (95% CI: 60.9-91.1%) for liver and bile duct sites (Figure 4) and Table 3. Stage distributions for each organ type are provided in Supplementary Table 3.

**Figure 4.**
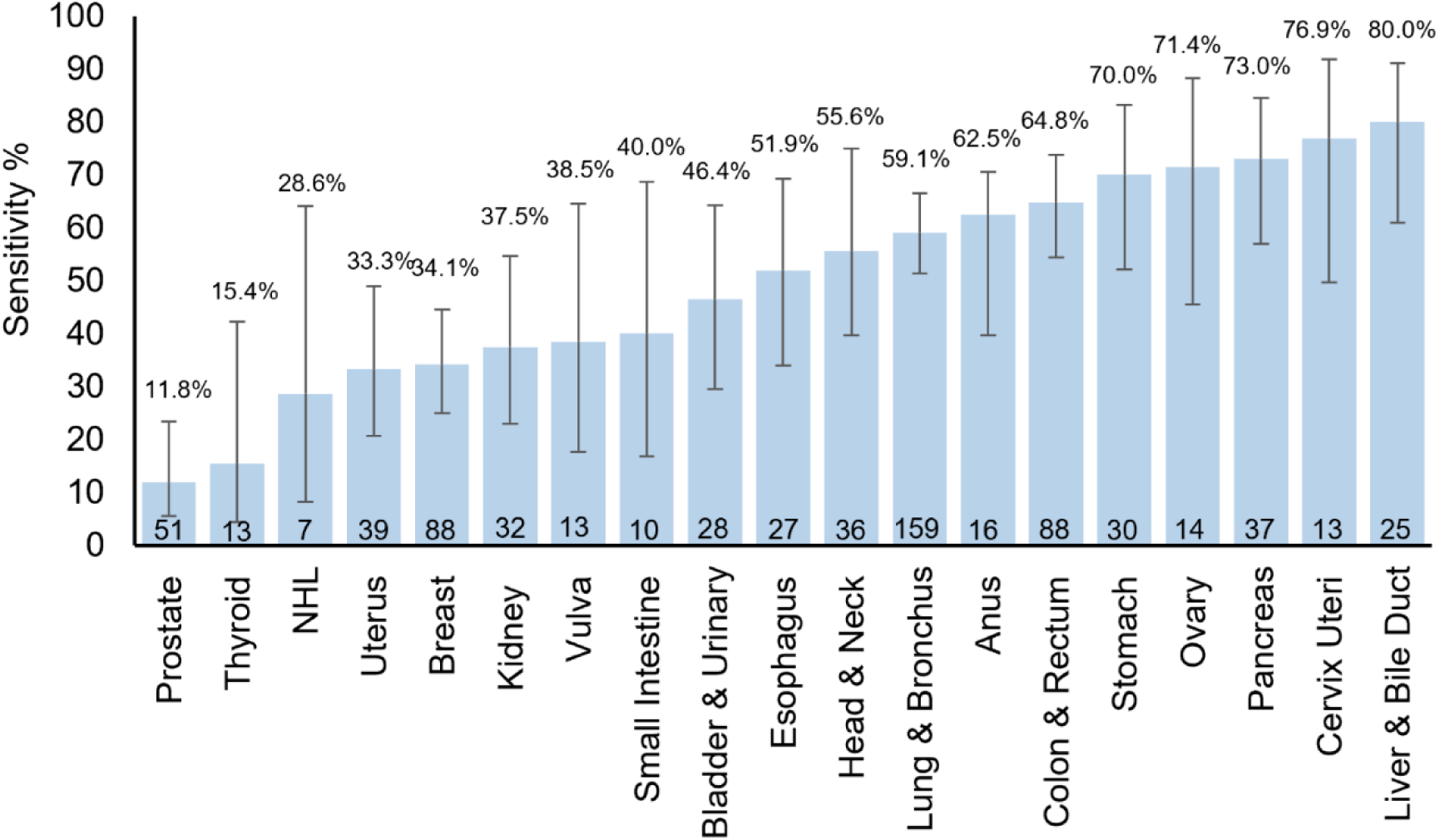
MP classifier organ-specific performance at 98.5% specificity*. MP, methylation-protein. NHL, Non-Hodgkin Lymphoma. *Multiple Myeloma (n=1) and testicular cancers (n=2) were omitted from the graph due to the small number of cases. The numbers at the base of each bar represent the number of cases.

**Figure 5.**
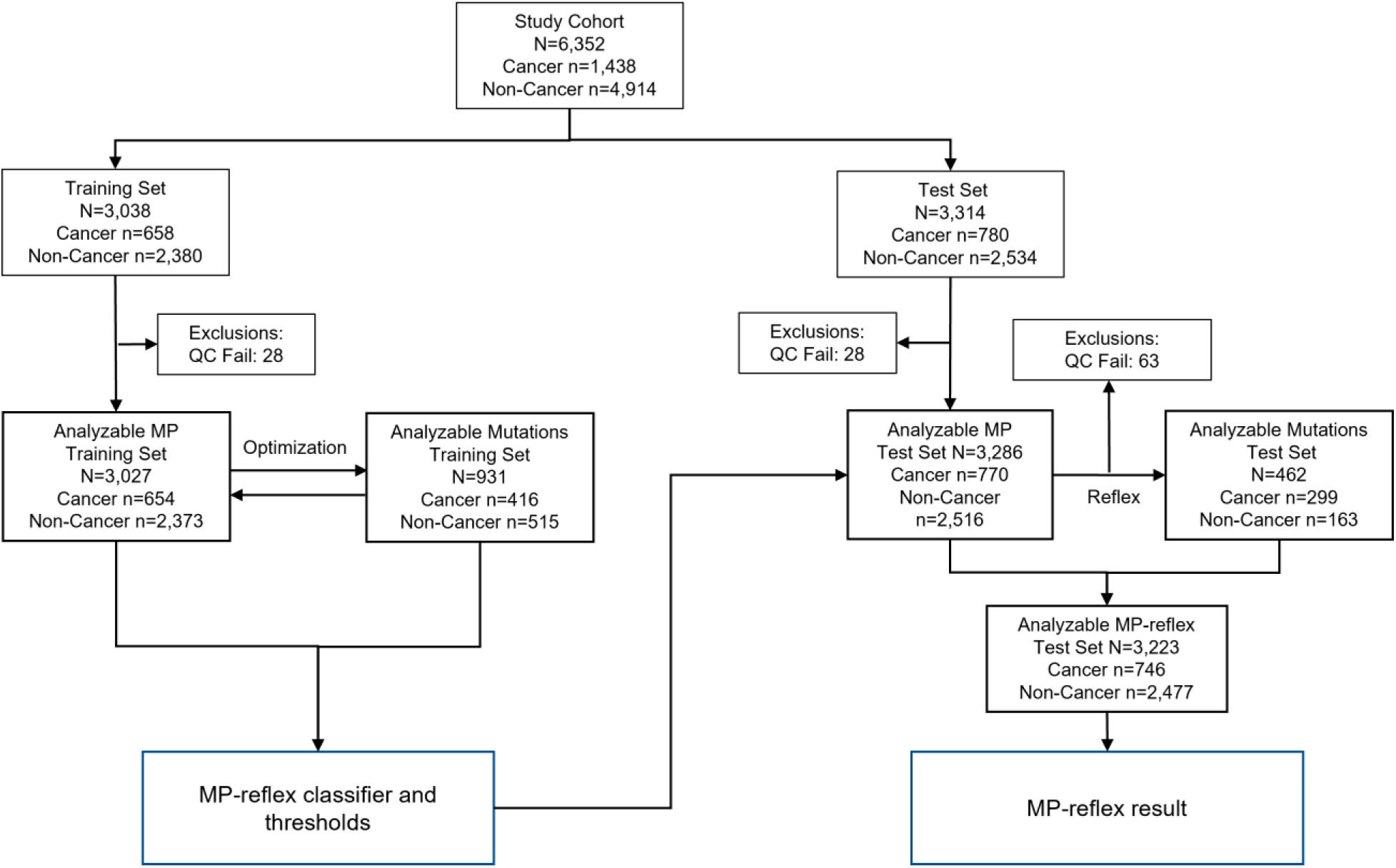
MP-reflex classifier development and test cohorts. MP, methylation-protein; QC, quality control.

**Table 2.**
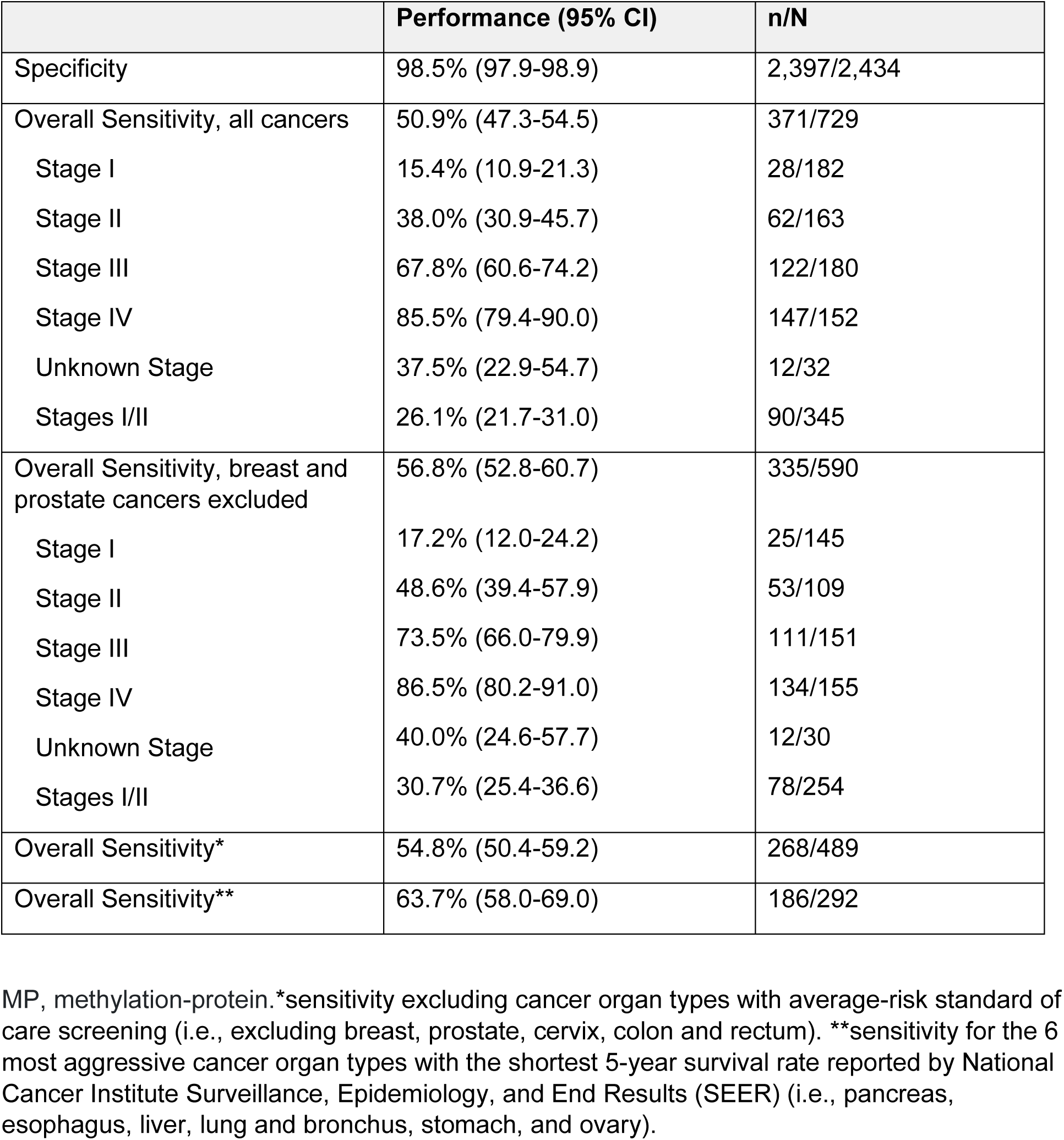
MP classifier test set performance.

|  | Performance (95% CI) | n/N |
| --- | --- | --- |
| Specificity | 98.5% (97.9-98.9) | 2,397/2,434 |
| Overall Sensitivity, all cancers | 50.9% (47.3-54.5) | 371/729 |
| Stage I | 15.4% (10.9-21.3) | 28/182 |
| Stage II | 38.0% (30.9-45.7) | 62/163 |
| Stage III | 67.8% (60.6-74.2) | 122/180 |
| Stage IV | 85.5% (79.4-90.0) | 147/152 |
| Unknown Stage | 37.5% (22.9-54.7) | 12/32 |
| Stages I/II | 26.1% (21.7-31.0) | 90/345 |
| Overall Sensitivity, breast and prostate cancers excluded | 56.8% (52.8-60.7) | 335/590 |
| Stage I | 17.2% (12.0-24.2) | 25/145 |
| Stage II | 48.6% (39.4-57.9) | 53/109 |
| Stage III | 73.5% (66.0-79.9) | 111/151 |
| Stage IV | 86.5% (80.2-91.0) | 134/155 |
| Unknown Stage | 40.0% (24.6-57.7) | 12/30 |
| Stages I/II | 30.7% (25.4-36.6) | 78/254 |
| Overall Sensitivity* | 54.8% (50.4-59.2) | 268/489 |
| Overall Sensitivity** | 63.7% (58.0-69.0) | 186/292 |

MP, methylation-protein.\*sensitivity excluding cancer organ types with average-risk standard of care screening (i.e., excluding breast, prostate, cervix, colon and rectum). \*\*sensitivity for the 6 most aggressive cancer organ types with the shortest 5-year survival rate reported by National Cancer Institute Surveillance, Epidemiology, and End Results (SEER) (i.e., pancreas, esophagus, liver, lung and bronchus, stomach, and ovary).

**Table 3.** MP classifier test set sensitivity by organ type and stage.

| Organ Type | Sensitivity (95% Confidence Interval) |  |  |  |  |
| --- | --- | --- | --- | --- | --- |
|  | Stage I | Stage II | Stage III | Stage IV | Unknown Stage |
| Lung and Bronchus (n=159) | 13%<br>(6.1%, 25.7%)<br>n=46 | 52.2%<br>(33%, 70.8%)<br>n=23 | 77.1%<br>(63.5%, 86.7%)<br>n=48 | 92.3%<br>(79.7%, 97.3%)<br>n=39 | 100%<br>(43.9%, 100%)<br>n=3 |
| Breast (n=88) | 8.3%<br>(2.3%, 25.8%)<br>n=24 | 30%<br>(16.7%, 47.9%)<br>n=30 | 47.8%<br>(29.2%, 67%)<br>n=23 | 88.9%<br>(56.5%, 98%)<br>n=9 | 0%<br>(0%, 65.8%)<br>n=2 |
| Colon and Rectum (n=88) | 15.4%<br>(4.3%, 42.2%)<br>n=13 | 50%<br>(29%, 71%)<br>n=18 | 76%<br>(56.6%, 88.5%)<br>n=25 | 92.6%<br>(76.6%, 97.9%)<br>n=27 | 40%<br>(11.8%, 76.9%)<br>n=5 |
| Prostate (n=51) | 7.7%<br>(1.4%, 33.3%)<br>n=13 | 0%<br>(0%, 13.8%)<br>n=24 | 0%<br>(0%, 39%)<br>n=6 | 62.5%<br>(30.6%, 86.3%)<br>n=8 |  |
| Uterus (n=39) | 15%<br>(5.2%, 36%)<br>n=20 | 25%<br>(4.6%, 69.9%)<br>n=4 | 57.1%<br>(25%, 84.2%)<br>n=7 | 83.3%<br>(43.6%, 97%)<br>n=6 | 0%<br>(0%, 65.8%)<br>n=2 |
| Pancreas (n=37) | 42.9%<br>(15.8%, 75%)<br>n=7 | 50%<br>(21.5%, 78.5%)<br>n=8 | 100%<br>(64.6%, 100%)<br>n=7 | 92.9%<br>(68.5%, 98.7%)<br>n=14 | 0%<br>(0%, 79.4%)<br>n=1 |
| Head and Neck (n=36) | 40%<br>(16.8%, 68.7%)<br>n=10 | 40%<br>(11.8%, 76.9%)<br>n=5 | 54.5%<br>(28%, 78.7%)<br>n=11 | 80%<br>(49%, 94.3%)<br>n=10 |  |
| Kidney (n=32) | 0%<br>(0%, 29.9%)<br>n=9 | 50%<br>(15%, 85%)<br>n=4 | 25%<br>(4.6%, 69.9%)<br>n=4 | 81.8%<br>(52.3%, 94.9%)<br>n=11 | 0%<br>(0%, 49%)<br>n=4 |
| Stomach (n=30) | 20%<br>(3.6%, 62.4%)<br>n=5 | 57.1%<br>(25%, 84.2%)<br>n=7 | 85.7%<br>(48.7%, 97.4%)<br>n=7 | 100%<br>(70.1%, 100%)<br>n=0 | 50%<br>(9.5%, 90.5%)<br>n=2 |
| Bladder and Urinary<br>(n=28) | 14.3%<br>(2.6%, 51.3%)<br>n=7 | 44.4%<br>(18.9%, 73.3%)<br>n=9 | 75%<br>(30.1%, 95.4%)<br>n=4 | 66.7%<br>(30%, 90.3%)<br>n=6 | 50%<br>(9.5%, 90.5%)<br>n=2 |
| Esophagus<br>(n=27) | 0%<br>(0%, 39%)<br>n=6 | 20%<br>(3.6%, 62.4%)<br>n=5 | 75%<br>(40.9%, 92.9%)<br>n=8 | 85.7%<br>(48.7%, 97.4%)<br>n=7 | 100%<br>(20.7%, 100%)<br>n=1 |
| Liver and Bile Duct (n=25) | 40%<br>(11.8%, 76.9%)<br>n=5 | 83.3%<br>(43.6%, 97%)<br>n=6 | 100%<br>(67.6%, 100%)<br>n=8 | 100%<br>(56.6%, 100%)<br>n=5 | 0%<br>(0%, 79.4%)<br>n=1 |
| Anus (n=16) | 0%<br>(0%, 79.4%)<br>n=1 | 62.5%<br>(30.6%, 86.3%)<br>n=8 | 66.7%<br>(20.8%, 93.9%)<br>n=3 | 100%<br>(43.9%, 100%)<br>n=3 | 0%<br>(0%, 79.4%)<br>n=1 |
| Ovary (n=14) | 0%<br>(0%, 79.4%)<br>n=1 | 0%<br>(0%, 79.4%)<br>n=1 | 85.7%<br>(48.7%, 97.4%)<br>n=7 | 66.7%<br>(20.8%, 93.9%)<br>n=3 | 100%<br>(34.2%, 100%)<br>n=2 |
| Vulva (n=13) | 0%<br>(0%, 39%)<br>n=6 |  | 66.7%<br>(20.8%, 93.9%)<br>n=3 | 100%<br>(34.2%, 100%)<br>n=2 | 50%<br>(9.5%, 90.5%)<br>n=2 |
| Cervix Uteri<br>(n=13) | 100%<br>(34.2%, 100%)<br>n=2 | 66.7%<br>(20.8%, 93.9%)<br>n=3 | 100%<br>(43.9%, 100%)<br>n=3 | 100%<br>(34.2%, 100%)<br>n=2 | 33.3%<br>(6.1%, 79.2%)<br>n=3 |
| Thyroid (n=13) | 0%<br>(0%, 49%)<br>n=4 | 0%<br>(0%, 49%)<br>n=4 |  | 50%<br>(15%, 85%)<br>n=4 | 0%<br>(0%, 79.4%)<br>n=1 |
| Small Intestine<br>(n=10) |  | 66.7%<br>(20.8%, 93.9%)<br>n=3 | 25%<br>(4.6%, 69.9%)<br>n=4 | 33.3%<br>(6.1%, 79.2%)<br>n=3 |  |
| Non-Hodgkin Lymphoma (n=7) | 0%<br>(0%, 79.4%)<br>n=1 |  | 0%<br>(0%, 65.8%)<br>n=2 | 50%<br>(15%, 85%)<br>n=4 |  |
| Testis (n=2) | 50%<br>(9.5%, 90.5%)<br>n=2 |  |  |  |  |
| Multiple Myeloma<br>(n=1) |  | 0%<br>(0%, 79.4%)<br>n=1 |  |  |  |

MP, methylation-protein.

MP test set performance from all enrollment sites was also compared to the performance from sites that were unique in the test set to determine if collection site influenced the classifier performance. Thirty-one percent of test set samples (non-cancers n=597; cancers n=383) were obtained from unique collection sites (n=71 sites) that were not included in the training set. The observed unique test set collection site overall sensitivity of 56.7% (95% CI: 51.7-61.5%) and 99.3% (95% CI: 98.3-99.7%) specificity were similar to the overall sensitivity and specificity observed for the full test set (sensitivity: 50.9% (95% CI: 47.3-54.5%); specificity 98.5% (95% CI: 97.9-98.9). Performance by stage was also similar between these test set cohorts (Supplementary Figure 4).

### Methylation Protein (MP)-reflex classifier training and testing

#### MP-reflex classifier training

6,352 samples (1,438 cancers; 4,914 non-cancers) were utilized for the MP-reflex classifier development and testing as outlined in Figure 5. Participant demographic information for the analyzable MP-reflex classifier training and test sets is shown in Supplementary Table 4. Of the 3,038 samples pre-assigned to the training set, 3,027 analyzable samples (654 cancers; 2,373 non-cancers) were used to train the MP classifier as outlined above as well as for setting the lower bound classification between high-confidence negative and samples with middle zone results. The mutation classifier was trained on a subset of 931 (416 cancers and 515 non-cancers).

### MP-reflex classifier test set performance

3,223 analyzable samples (746 cancers; 2,477 non-cancers) representing 20 organ types (stomach, ovary, prostate, colon and rectum, lung and bronchus, bladder and urinary, breast, head and neck (squamous cell), kidney, pancreas, uterus, esophagus, anus, vulva, liver and bile duct, small intestine, thyroid, cervix uteri, testis, Non-Hodgkin Lymphoma) and all stages were used to test the MP-reflex classifier (Figure 5). Multiple Myeloma was excluded from the analysis due to a small sample number (n=1). MP classifier resulted in 462 (299 cancers; 163 non-cancers) samples that fell into a “middle zone” (i.e., MP classifier results that fell in-between confident positive and confident negative calls), and these samples were analyzed using the mutations classifier.

By applying the tuned mutations thresholds, the MP-reflex classifier demonstrated a specificity of 98.5% (95% CI: 97.9-98.9%), with an overall sensitivity of 55.2% (95% CI: 51.6-58.8%), and sensitivities of 19.0% (95% CI: 14.0-25.3%), 29.5% (95% CI: 25.0-34.5%), 41.2% (95% CI: 34.0-48.8%), 71.9% (95% CI: 65.0-77.9%), 89.4% (95% CI: 84.1-93.1%) and 46.9% (95% CI: 30.9-63.6%) for stages I, I and II combined, II, III, IV, and unknown, respectively for all 20 organ types. The MP-reflex classifier performance was also assessed with breast and prostate cancers excluded, with an observed overall sensitivity of 61.6% (95% CI: 57.6-65.3%), and sensitivities of 21.9% (95% CI: 16.0-29.3%), 35.2% (95% CI: 29.6-41.2%), 52.7% (95% CI: 43.5-61.8%), 75.9% (95% CI: 68.7-81.9%), 90.8% (95% CI: 85.4-94.3%) and 51.7% (95% CI: 34.4-68.6%) for stages I, I and II combined, II, III, IV, and unknown, respectively. Overall sensitivity excluding cancer organ types with average-risk SoC screening (i.e. excluding breast, prostate, cervix, colon and rectum) was 59.9% (95% CI: 55.1-63.6%) and 66.9% (95% CI: 61.4-71.9%) for the six most aggressive cancer organ types with the shortest 5-year survival rate^1^ (i.e. pancreas, esophagus, liver, lung and bronchus, stomach, and ovary) (Table 4).

**Table 4.** MP-reflex classifier test set performance.

| Test Set | Estimate % (95% CI) | n/N |
| --- | --- | --- |
| **Specificity** | 98.5% (97.9-98.9) | 2,440/2,477 |
| **Sensitivity, all cancers** | 55.2% (51.6-58.8) | 412/746 |
| Stage I | 19.0% (14.0-25.3) | 35/184 |
| Stage II | 41.2% (34.0-48.8) | 68/165 |
| Stage III | 71.9% (65.0-77.9) | 133/185 |
| Stage IV | 89.4% (84.1-93.1) | 161/180 |
| Unknown stage | 46.9% (30.9-63.6) | 15/32 |
| Stage I, II | 29.5% (25.0-34.5) | 103/349 |
| Stage I-III | 44.2% (40.0-48.4) | 236/534 |
| **Sensitivity-excluding breast and prostate** | 61.6% (57.6-65.3) | 373/606 |
| Stage I | 21.9% (16.0-29.3) | 32/146 |
| Stage II | 52.7% (43.5-61.8) | 58/110 |
| Stage III | 75.9% (68.7-81.9) | 120/158 |
| Stage IV | 90.8% (85.4-94.3) | 148/163 |
| Unknown stage | 51.7% (34.4-68.6) | 15/29 |
| Stage I, II | 35.2% (29.6-41.2) | 90/256 |
| Stage I-III | 50.7% (45.9-55.5) | 210/414 |
| **Sensitivity\*** | 59.4% (55.1-63.6) | 300/505 |
| **Sensitivity\*\*** | 66.9% (61.4-71.9) | 206/308 |

MP, methylation-protein.\*sensitivity excluding cancer organ types with average-risk standard of care screening (i.e., excluding breast, prostate, cervix, colon and rectum). \*\*sensitivity for the 6 most aggressive cancer organ types with the shortest 5-year survival rate reported by National Cancer Institute Surveillance, Epidemiology, and End Results (SEER) (i.e., pancreas, esophagus, liver, lung and bronchus, stomach, and ovary).

### MP and MP-reflex classifier performance comparison

MP and MP-reflex classifier performance were compared using the MP-reflex test set (746 cancers and 2,477 non-cancers). At specificities of ≥98.5%, the MP-reflex classifier showed absolute sensitivity (absolute change%=MP-reflex sensitivity% minus MP sensitivity%) increases of 3.8%, 3.1%, 2.4%, 1.1%, and 2.7%, for stages I, I and II combined, II, III, and IV, respectively, compared to the MP classifier. The overall absolute sensitivity gain of the MP-reflex classifier was 2.8% (5.3% relative gain; defined as MP-reflex sensitivity% minus MP sensitivity% divided by MP sensitivity% x 100) compared to the MP classifier sensitivity. Notably, the absolute sensitivity gain of the MP-reflex classifier for early stages (stages I and II) was 3.1% (11.7% relative gain) compared to the MP classifier (Figure 6). The relative gain in sensitivity was greatest in stage I disease at 25.0%.

**Figure 6.**
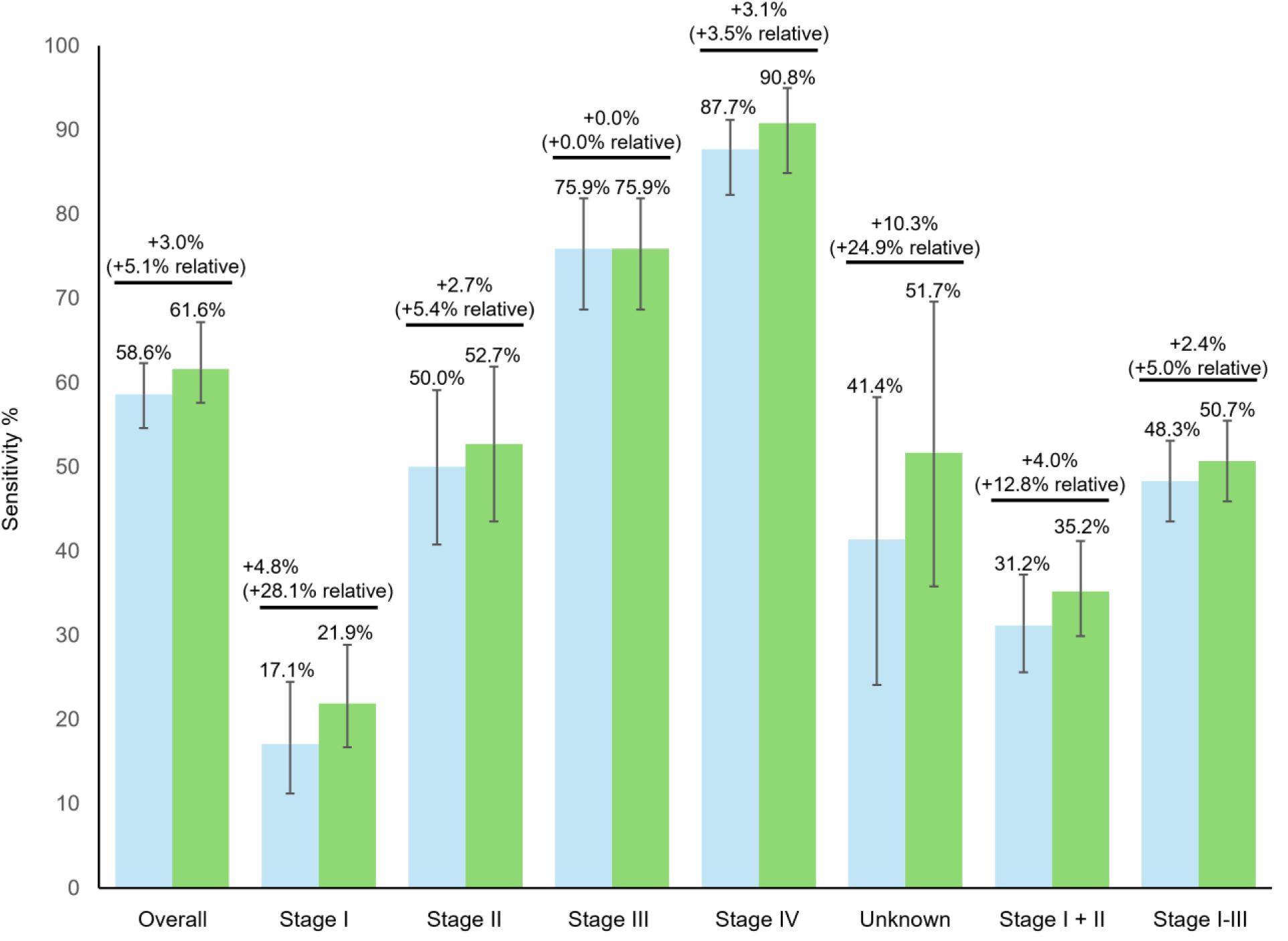
MP and MP-reflex classifier performance comparison (breast and prostate cancers excluded)* MP, methylation-protein.*using MP-reflex test set cohort (746 cancers and 2,477 non-cancers). The absolute change=%MP reflex sensitivity-%MP sensitivity. The relative change=%MP reflex sensitivity-%MP sensitivity/%MP sensitivity.

MP and MP-reflex classifier performance was also compared for cancer organ types with average-risk standard of care screening (i.e., excluding breast, prostate, cervix, colon and rectum). The 59.4% (95% CI: 55.1-63.6%) observed overall sensitivity for the MP-reflex classifier represents a 2.8% absolute increase compared to the MP classifier (56.6%, 95% CI: 52.3-60.9%). Additionally, the 66.9% (95% CI: 61.4-71.9%) overall sensitivity observed for the MP-reflex classifier represents a 2.0% absolute increase for the six most aggressive cancer organ types (pancreatic, esophagus, liver, lung, stomach, and ovarian) compared to the MP classifier (64.9%, 95% CI: 59.5-70.1%). The relative sensitivity gain was 23.0% for stage I and 4.6% for stage II in this subgroup.

## Discussion

Building upon our previous efforts in MCED assay development, we evaluated the performance of two different multi-target biomarker configurations in a large, multi-center prospectively collected case-control study. The ASCEND-2 study included a racially, ethnically, and geographically diverse sample representative of the U.S. population.^35^ Age, sex, and race/ethnicity were comparable between the cancer and non-cancer groups in both the training and test sets. Organ sites and cancer stages (which were approximately equally distributed) collectively represent most incident cancers observed in the U.S. population, including aggressive cancers and cancers without guideline-recommended screening options.

We have previously reported case-control study feasibility data for three (methylation, protein, aneuploidy) and four biomarker classes (methylation, protein, aneuploidy, and mutations) MCED assay configurations across fifteen organ sites. In this early detection biomarker feasibility work, the observed overall sensitivities were 53.4% and 61.0% at specificities of 98.8% and 98.2% for the combined three or four biomarker classes, respectively.^25^ We then refined and optimized each biomarker algorithm as well as the overarching cancer classifier, resulting in improved overall sensitivities of 55.2% and 62.4% at specificities of 99.0% and 98.0% for the combined three or four biomarker classes, respectively.^26^

Based on individual biomarker contributions to overall performance, in this study we initially focused on a combined MP assay configuration in an ongoing effort to increase assay efficiency while maintaining sensitivity and specificity. The MP assay classifier demonstrated an overall sensitivity of 50.9%, which was comparable to the overall sensitivities reported above for the three biomarkers (methylation, protein, aneuploidy) MCED assay configuration while maintaining a high specificity of 98.5%. Measuring cfDNA-based biomarkers for breast and prostate cancers is particularly challenging due to nominal circulating tumor DNA shedding rates associated with low tumor burden and high variability in circulating tumor DNA due to tumor heterogeneity.^37–39^ Further, these two cancers have established high-performing screening options. The overall sensitivity of MP performance with breast and prostate cancers excluded was 56.8%.

MCED tests currently in development are intended to complement, not replace, SoC cancer screening modalities. Interestingly, when cancer organ types with average-risk SoC screening were excluded except for lung cancers, which do not have a screening option for the general population (i.e., excluding breast, prostate, cervix, colon and rectum), the MP classifier demonstrated an overall sensitivity of almost 55%. In addition, overall sensitivity was nearly 64% for the six most aggressive cancer types with the shortest 5-year survival. Although preliminary, these data suggest that MCED tests may broaden population wide screening availability for individuals who are not adherent with current screening programs and for those with cancers without SoC screening options.

To further enhance cancer detection, particularly for stage I and II cancers, without substantially increasing testing complexity and cost, a second three-biomarker class approach was also explored. In this approach, methylation and protein testing are performed on all samples first followed by mutation testing for a fraction of samples that were neither highly positive nor negative for the methylation and protein biomarkers. By interrogating the mutation profiles of tumor hotspots for MP “middle zone” samples, the MP-reflex approach demonstrated a sensitivity increase of 3.1% (11.7% relative gain) for stage I and II cancers while maintaining a specificity of 98.5%. The greatest relative improvement in sensitivity was observed for the detection of stage I cancers. While we achieved modest improvements for stage I and II detection with the reflex approach; early-stage sensitivities were lower than those observed for recommended single cancer screening tests that directly interrogate the organ of interest at lower specificities. Sensitivity improvements were also observed when cancer types with average risk SoC screening were considered (2.8% absolute increase for MP-reflex, data not shown) and for the six most aggressive cancer types (3.0% absolute increase for MP-reflex).

In addition to the enhanced performance exhibited by the MP-reflex classifier for early-stage cancer detection, the reflex methodology is also more cost-effective compared to conducting methylation, protein, and mutation testing simultaneously on all samples. The MP classifier result triggered the reflex mutations testing in about 16% of the test set samples, achieving a favorable balance between improved early-stage sensitivity and reduced assay cost. Reduction in assay cost is critical to support the ability to deliver a widely accessible assay for all those who might benefit from MCED screening.

## Limitations

Case-control studies like ASCEND-2 are an efficient data source for classifier training and assessing classifier performance in an independent hold-out test cohort. As with any case-control study, the approach applied here has limitations. First, ASCEND-2 enrollment criteria for the non-cancer control cohort depended on self-reported and recorded information to define cancer-free status just prior to or at the time of enrollment. Full adherence with recommended cancer screening was not required. Also, ASCEND-2 did not have a pre-defined built in longitudinal follow up included that would allow us to identify any cancer that developed within a one-to-two-year timeframe from the time of the blood draw. Therefore, the training and test cohorts likely included unobserved cancer or pre-cancers that could have resulted in a cancer-related biomarker signal. Approximately 1% of individuals above the age of 50 (almost 1.8 million individuals estimated in 2025) are diagnosed with cancer each year.^40^ Thus, undiagnosed cancers are expected in the general population. For screening tests with a high specificity that aim for no more than 1.5% false positive results, this level of undiagnosed cancer burden represents a challenge. This would have complicated the training process as well as the interpretation of the testing results. One approach in the future is to further evaluate these results by utilization of privacy-preserving record linkage (PPRLS) techniques (i.e., tokenization) to determine subsequent cancer status by linking participant data sets from electronic medical records with results from the non-cancer test cohort.

Second, the enrollment of cancer participants in ASCEND-2 was not limited to a single diagnostic pathway that would be reflective of an asymptomatic screening test user. Specifically, the ASCEND-2 cancer cohort participants were identified by one of these mechanisms: screening, incidental findings, or by symptoms. It is expected, depending on this diagnostic enrollment mechanism, that there are differences in biomarker signal strength and distribution. Depending on the cancer type and stage, the ratio between these three pathways could fluctuate and may not reflect the diagnostic pathways in a screening setting.

ASCEND-2 study enrollment was enriched for lung and bronchus cancer, while breast, prostate, and hematological cancers were under-represented. This may have influenced performance results as sensitivities vary widely across individual cancer types. Disproportionate over-sampling of specific cancer types with higher sensitivities (lung and bronchus) while under sampling those with lower sensitivities (breast, prostate) would be expected to result in higher overall sensitivities. In addition, we targeted equal stage distribution across all the cancer types included in ASCEND-2. Consequently, compared to SEER incidence, a higher number of later-stage cancers were included in the study.

Third, the prospectively collected ASCEND-2 included a diverse population; however, the analyses were performed in a case-control fashion without the return of results to participants. Reported MCED test sensitivities have been higher for case–control studies (70.0% overall sensitivity for Cohen’s CancerSEEK case–control study) compared to prospective studies (27.1% overall sensitivity) observed in DETECT-A.^24, 27^ Prospective clinical validation studies in average risk individuals will reflect assay and clinical diagnostic work-up performance in the intended-use population.

Finally, reporting of test performance in subsets of the study population defined by age, gender, race, ethnicity, risk factors for cancer and other factors of interest is beyond the scope of this report owing the small sample size available for such analyses. Future research will be needed to further evaluate test performance in subsets of the overall population.

## Conclusion

MP and MP-reflex classifier strategies resulted in overall sensitivities of 50.9% and 55.2% for a broad range of cancers respectively, at a specificity of 98.5%. These results represent a progress report from an ongoing program developing a multi-biomarker class MCED test.

A real-world evidence registry, Falcon (Clinical Trial Identifier: NCT06589310) was initiated in 2024 to prospectively investigate test performance and the uptake, diagnostic journey, adherence with guideline-recommended cancer screening, outcomes, and psychological impacts of MCED testing.

## Data Availability

All data produced in the present study are available upon reasonable request to the authors.

## Acknowledgements

The authors would like to thank the following individuals: Jing Tong, Amira Best, David Sims, Jeremy Thorpe, Christopher Gault, Farhad Hatami and Mark Evans for feedback on study design and execution, sample processing and data preparation, Natsuyo Aoyama for project management support, Fikrullah Kisa, Peter Ngam, Jerry Machado and Raja Kakuturu for wet laboratory leadership, Amy Lehman for statistical analysis and Amin Mazloom (Exact Sciences Corporation, Madison, WI) for leadership and feedback. Carolyn Hall and Feyza Sancar (Exact Sciences Corporation, Madison, WI) provided manuscript writing and editorial support. The authors would also like to thank the patients who generously participated in the original sample collection studies and the principal investigators and institutions who oversaw their enrollment.

## Author Contributions

Conceptualization: Frank Diehl

Methodology: Vladimir Gainullin, Frank Diehl

Formal Analysis: Vladimir Gainullin, Mael Manesse, Fanglei Zhuang, Melissa Gray, Hee Jung Hwang, Gustavo C. Cerqueira, Christopher L. Nobles, Madhav Kumar, Kevin Arvai, Xi Chen, Christopher Tyson, Chen Ji, Jin Bae, Violeta Beleva Guthrie, Jaspreet Kaur, Viatcheslav E. Katerov, Larson Hogstrom, Leonardo Hagmann, Philip J. Uren, Vuna Fa, Abigail McElhinny

Investigation: Gerard Silvestri, David Chesla, Robert Given

Writing-Original Draft: Vladimir Gainullin, Frank Diehl, Tomasz M. Beer

Writing-Review and Editing: Vladimir Gainullin, Mael Manesse, Fanglei Zhuang, Melissa Gray, Hee Jung Hwang, Gustavo C. Cerqueira, Christopher L. Nobles, Madhav, Kumar, Kevin Arvai, Xi Chen, Chris Tyson, Chen Ji, Jin Bae, Violeta Beleva Guthrie, Jaspreet Kaur, Viatcheslav E. Katerov, Larson Hogstrom, Leonardo Hagmann, Philip Uren, Jorge Garces, Hatim T. Allawi, Vuna Fa, Abigail McElhinny, Gerard Silvestri, David Chesla, Robert Given, Tomasz M. Beer, Frank Diehl Project Administration: Jorge Garces, Hatim T. Allawi, Tomasz M. Beer, Frank Diehl

## Abbreviations

ASCEND-2: Ascertaining Serial Cancer patients to Enable New Diagnostic 2
cfDNA: cell-free DNA
CHIP: clonal hematopoiesis of indeterminate potential
CV: cross-validation
DETECT-A: <u>D</u>etecting cancers <u>E</u>arlier <u>T</u>hrough <u>E</u>lective mutation-based blood <u>C</u>ollection and <u>T</u>esting
ECL: electrochemiluminescence
gDNA: genomic DNA
LQAS: Long-probe Quantitative Amplified Signal
MCED: multi-cancer early detection
MDM: methylated DNA marker
MP: methylation and protein
MP-reflex: methylation and protein plus somatic mutation reflex
PCR: polymerase chain reaction
PPRLS: privacy-preserving record linkage
QC: quality control
ROC AUC: Receiver Operating Characteristic Area Under the Curve
SEER: National Cancer Institute Surveillance, Epidemiology, and End Results
SoC: standard of care
TELQAS: Target Enrichment Long-probe Quantitative Amplified Signal
USPSTF: United States Preventive Services Taskforce

