## Supplementary material for "Performance of a multi-target, multi-cancer early detection (MCED) blood test in a prospectively collected cohort": MedRxiv Supplementary Materials 8-24-2025.docx

*Former employee of Exact Sciences Corporation

Supplementary Table 1. Total cancer study cohort (n=1,438) by organ type compared to normalized SEER incidence

| **Cancer Organ Type** | **% of total (number of samples)** | **Normalized SEER Incidence %^a^** |
| --- | --- | --- |
| anus | 1.9% (28) | 0.6% |
| bladder and urinary | 4.5% (65) | 3.0% |
| breast^b^ | 11.5% (165) | 17.7% |
| cervix uteri | 1.5% (22) | 0.6% |
| colon and rectum | 11.2% (161) | 9.9% |
| esophagus | 3.7% (53) | 1.3% |
| head and neck | 5.4% (78) | 4.3% |
| kidney | 5.6% (80) | 4.6% |
| liver and bile duct | 3.5% (50) | 2.9% |
| lung and bronchus^c^ | 24.2% (348) | 15.8% |
| lymphatic system, multiple myeloma^d^ | 0.1% (1) | 2.0% |
| lymphatic system, non-Hodgkin’s lymphoma^d^ | 0.6% (9) | 5.0% |
| ovary | 2.5% (37) | 1.4% |
| pancreas | 5.3% (76) | 3.8% |
| prostate^b^ | 4.6% (66) | 17.5% |
| small intestine | 0.8% (12) | 0.7% |
| stomach | 4.0% (58) | 1.9% |
| testis | 0.1% (2) | 0.1% |
| thyroid | 1.7% (25) | 2.3% |
| uterus | 5.6% (81) | 4.2% |
| vulva | 1.5% (21) | 0.4% |

^a^Normalized to 2023 National Cancer Institute Surveillance, Epidemiology, and End Results (SEER, accessed March 2024) proportions are adjusted to account for only cancer types included in this study. Values do not account for the incidence of seven rare cancer types included in the SEER database but not included in ASCEND 2. ^b^Breast and prostate selection was de-prioritized due to available screening options and low expected cfDNA shedding, respectively. ^c^Lung and bronchus cancer selection was slightly enriched because of potentially high clinical utility. ^d^Hematologic cancer cases were lower than SEER incidence rates due to a lower-than-expected enrollment rate.

Supplementary Table 2. Participant demographics of analyzable samples for MP classifier training and test sets with age and sample processing exclusions

| **Variable** | **Training Set**  **N=3,027** | | **Test Set**  **N=3,163** | |
| --- | --- | --- | --- | --- |
|  | **Non-Cancer**  **n=2,373** | **Cancer**  **n=654** | **Non-Cancer**  **n=2,434** | **Cancer**  **n=729** |
| **Sex, n (%)**  Female  Male | 1,344 (56.6%)  1,029 (43.4%) | 354 (54.1%)  300 (45.9%) | 1,392 (57.2%)  1,042 (42.8%) | 388 (53.2%)  341 (46.8%) |
| **Age, years**  Mean (SD)  (min, max) | 64.8 (8.0)  (50,96) | 66.9 (9.0)  (50,92) | 64.9 (7.8)  (50, 84) | 66.3 (8.3)  (50, 84) |
| **Race, n (%)**  White  Black or African American  Asian  American Indian/Alaska native  Hawaiian/Paciﬁc Islander  Mixed Race  Unknown/Missing | 1,540 (64.9%)  360 (15.2%)  84 (3.5%)  10 (0.4%)  3 (0.1%)  3 (0.1%)  14 (0.6%) | 560 (85.5%)  49 (7.5%)  24 (3.7%)  2 (0.3%)  0 (0.0%)  1 (0.2%)  18 (2.8%) | 2,009 (82.5%)  327 (13.4%)  55 (2.3%)  10 (0.4%)  2 (0.1%)  7 (0.3%)  24 (1.0%) | 587 (80.5%)  67 (9.2%)  31 (4.3%)  7 (1.0%)  1 (0.1%)  1 (0.1%)  35 (4.8%) |
| **Ethnicity, n (%)**  Not Hispanic/Latino  Hispanic/Latino  Unknown/Missing | 1,969 (83.0%)  390 (16.4%)  14 (0.6%) | 613 (93.7%)  25 (3.8%)  16 (2.4%) | 2,077 (85.3%)  341 (14.0%)  16 (0.7%) | 650 (89.2%)  49 (6.7%)  30 (4.1%) |

Supplementary Table 3. Stage distributions by organ type of the analyzable MP test set (n=729) with age and sample processing exclusions

| **Organ Type** | **Stage I** | **Stage II** | **Stage III** | **Stage IV** | **Unknown Stage** |
| --- | --- | --- | --- | --- | --- |
| Lung and Bronchus (n=159) | 46 | 23 | 48 | 39 | 3 |
| Breast (n=88) | 24 | 30 | 23 | 9 | 2 |
| Colon and Rectum (n=88) | 13 | 18 | 25 | 27 | 5 |
| Prostate (n=51) | 13 | 24 | 6 | 8 | 0 |
| Uterus (n=39) | 20 | 4 | 7 | 6 | 2 |
| Pancreas (n=37) | 7 | 8 | 7 | 14 | 1 |
| Head and Neck (n=36) | 10 | 5 | 11 | 10 | 0 |
| Kidney and Renal Pelvis (n=32) | 9 | 4 | 4 | 11 | 4 |
| Stomach (n=30) | 5 | 7 | 7 | 9 | 2 |
| Urinary Bladder (n=28) | 7 | 9 | 4 | 6 | 2 |
| Esophagus (n=27) | 6 | 5 | 8 | 7 | 1 |
| Liver and Bile Duct (n=25) | 5 | 6 | 8 | 5 | 1 |
| Anus (n=16) | 1 | 8 | 3 | 3 | 1 |
| Ovary (n=14) | 1 | 1 | 7 | 3 | 2 |
| Vulva (n=13) | 6 | 0 | 3 | 2 | 2 |
| Cervix Uteri (n=13) | 2 | 3 | 3 | 2 | 3 |
| Thyroid (n=13) | 4 | 4 | 0 | 4 | 1 |
| Small Intestine (n=10) | 0 | 3 | 4 | 3 | 0 |
| Non-Hodgkin Lymphoma (n=7) | 1 | 0 | 2 | 4 | 0 |
| Testis (n=2) | 2 | 0 | 0 | 0 | 0 |
| Multiple Myeloma (n=1) | 0 | 1 | 0 | 0 | 0 |
| Total | 182 | 163 | 180 | 172 | 32 |

Supplementary Table 4. Participant demographics of analyzable samples for MP-reflex classifier training and testing

|  | **MP-reflex Training Set**  **Cancer Non-Cancer Total**  **n=654 n=2,373 n=3,027** | | | **MP-reflex Test Set**  **Cancer Non-Cancer Total**  **n=746 n=2,477 n=3,223** | | |
| --- | --- | --- | --- | --- | --- | --- |
| **Sex n, (%)**    Female    Male | 354 (54.1%)    300 (45.9%) | 1,344 (56.6%)    1,029 (43.4%) | 1,698 (56.1%)    1,329 (43.9%) | 405 (54.3%)    341 (45.7%) | 1,423 (57.4%)    1,054 (42.6%) | 1,828 (56.7%)    1,395 (43.3%) |
| **Age years**    Mean (SD)    (Min,Max) | 66.9 (9.0)    (50, 92) | 64.8 (8.0)    (50, 96) | 65.3 (8.3)    (50, 96) | 66.9 (9.0)    (50, 92) | 65.2 (8.2)    (50, 101) | 65.6 (8.4)    (50, 101) |
| **Race n, (%)**    American Indian/Alaska Native    Asian    Black/African American    Mixed Race    Native Hawaiian/Other Pacific Islander  Unknown/Missing    White | 2 (0.3%)    24 (3.7%)    49 (7.5%)      1 (0.2%)      0 (0.0%)    18 (2.8%)    560 (85.6%) | 10 (0.4%)    84 (3.5%)    360 (15.2%)      3 (0.1%)      3 (0.1%)    14 (0.6%)    1,899 (80.0%) | 12 (0.4%)    108 (3.6%)    409 (13.5%)      4 (0.1%)      3 (0.1%)    32 (1.1%)    2,459 (81.2%) | 7 (0.9%)    26 (3.5%)    68 (9.1%)      1 (0.1%)      1 (0.1%)    37 (5.0%)    606 (81.2%) | 10 (0.4%)    58 (2.3%)    328 (13.2%)      8 (0.3%)      2 (0.1%)    24 (1.0%)    2,047 (82.6%) | 17 (0.5%)    84 (2.6%)    396 (12.3%)      9 (0.3%)      3 (0.1%)    61 (1.9%)    2,653 (82.3%) |
| **Ethnicity n, (%)**    Hispanic/Latino    Not Hispanic/Latino    Unknown/Missing | 25 (3.8%)    613 (97.3%)    16 (2.4%) | 390 (16.4%)    1,969 (83.0%)    14 (0.6%) | 415 (13.7%)    2,582 (85.3%)    30 (1.0%) | 52 (7.0%)    661 (6.9%)    33 (4.4%) | 342 (13.8%)    2,118 (85.5%)    17 (0.7%) | 394 (12.2%)    2,779 (86.2%)    50 (1.6%) |

Supplementary Table 5. MP performance estimation without age / sample processing exclusions

|  | **Performance (95% CI)** | **n/N** |
| --- | --- | --- |
| Specificity | 98.4% (97.9-98.9) | 2,477/2,516 |
| Overall Sensitivity, all cancers  Stage I  Stage II  Stage III  Stage IV  Unknown Stage  Stages I/II | 51.6% (48.0-55.1)  15.5% (11.1-21.2)  39.3% (32.2-46.8)  69.1% (62.2-75.2)  85.7% (79.9-90.1)  37.1% (23.2-53.7)  26.5% (22.2-31.3) | 397/770  30/194  66/168  132/191  156/182  13/35  96/362 |
| Overall Sensitivity, breast and prostate cancers excluded  Stage I  Stage II  Stage III  Stage IV  Unknown Stage  Stages I/II | 57.7% (53.8-61.5)  17.5% (12.3-24.3)  50.4% (41.4-59.5)  74.7% (67.5-80.8)  86.7% (80.6-91.0)  40.6% (24.6-57.7)  31.5% (26.2-37.3) | 361/626  27/154  57/113  121/162  143/165  13/32  84/267 |
| Overall Sensitivity* | 55.9% (51.6-60.1) | 291/521 |
| Overall Sensitivity** | 64.6% (59.2-69.7) | 201/311 |

*sensitivity excluding cancer organ types with average-risk standard of care screening (i.e., excluding breast, prostate, cervix, colon and rectum). **sensitivity for the 6 most aggressive cancer organ types with the shortest 5-year survival rate reported by National Cancer Institute Surveillance, Epidemiology, and End Results (SEER) (i.e., pancreas, esophagus, liver, lung and bronchus, stomach, and ovary).

Supplementary Figure 1. Comparison of sample counts between MP classifier training (left bar) and test (right bar) sets by organ site and cancer stage*

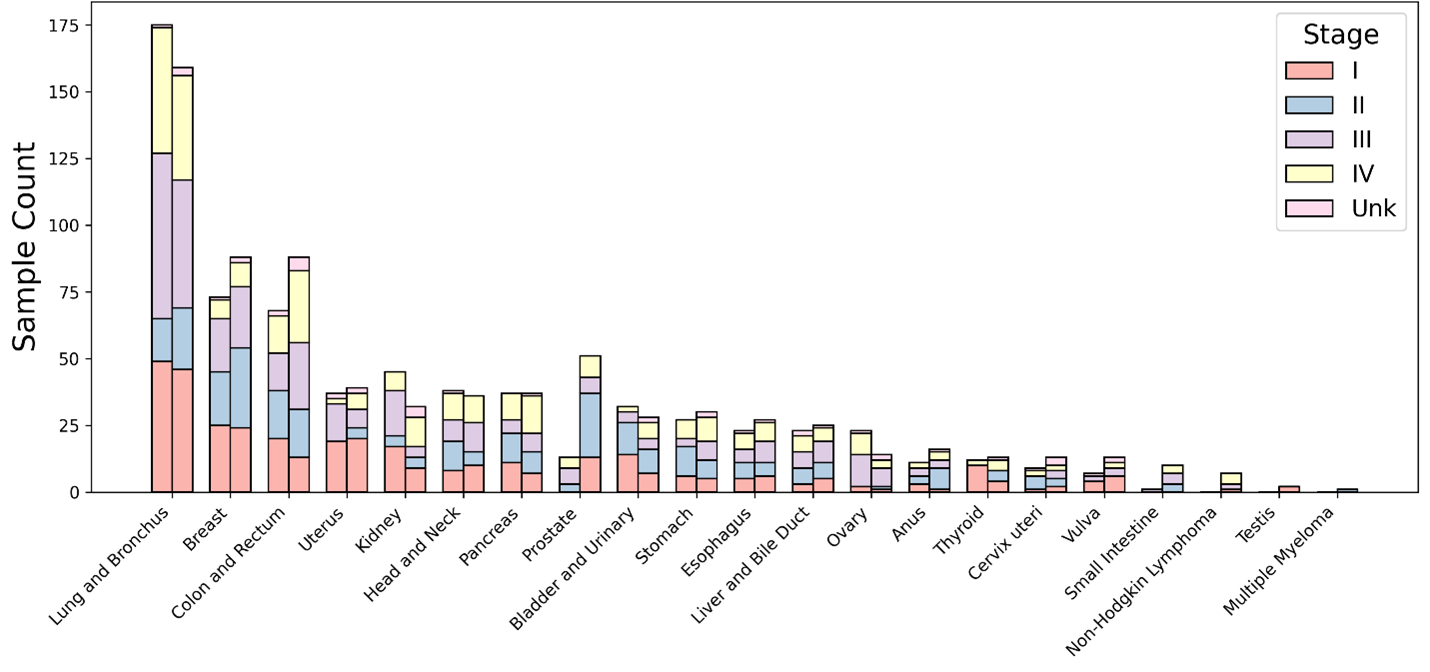

MP, methylation-protein.* left bar=training set, right bar = test set.

Supplementary Figure 2. Comparison of MP classifier performance by stage between the full training set (5x cross validation and mini-hold-out set; dark green), the 5x cross validation training set (light green) and test set (blue) performance comparison by stage

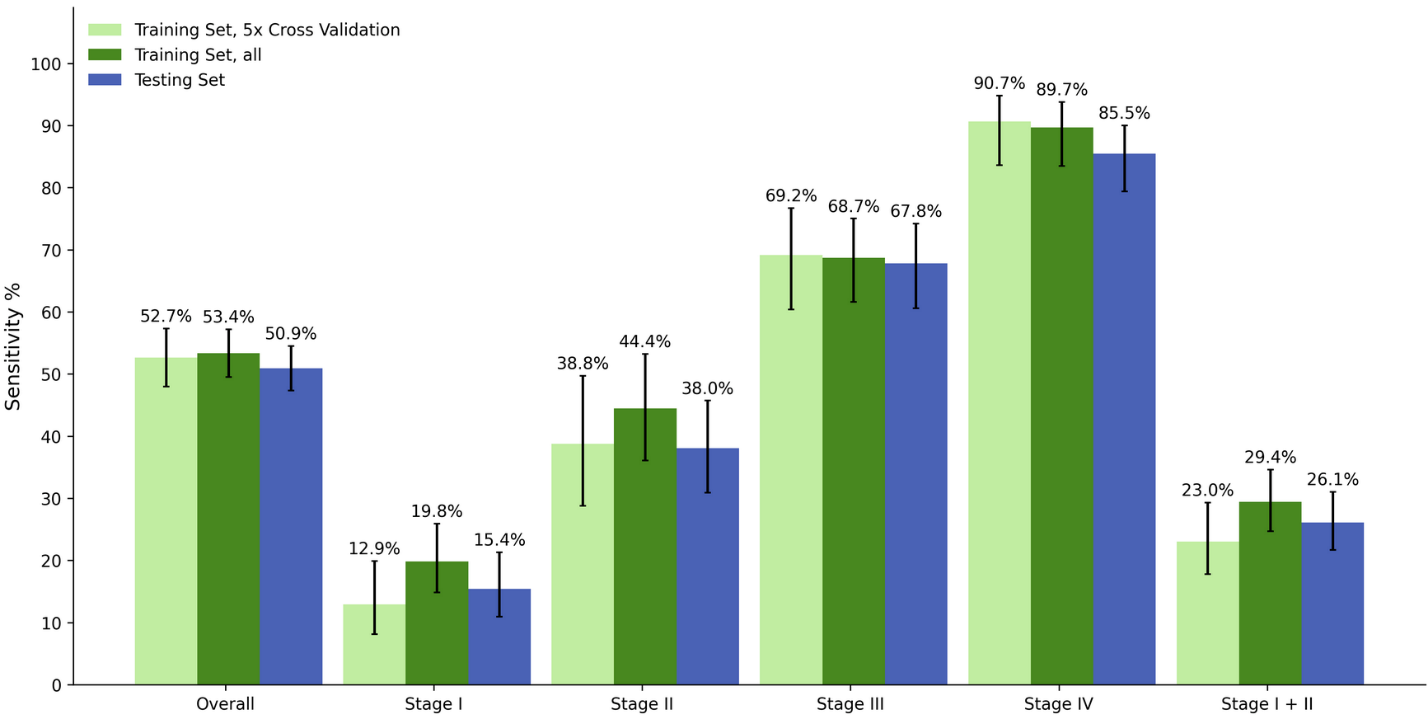

MP, methylation-protein.

Supplementary Figure 3. Comparison of MP classifier performance by tumor organ site between the full training set (5x cross validation and mini-hold-out set; dark green), the 5x cross validation training set (light green) and test set (blue) performance comparison by stage

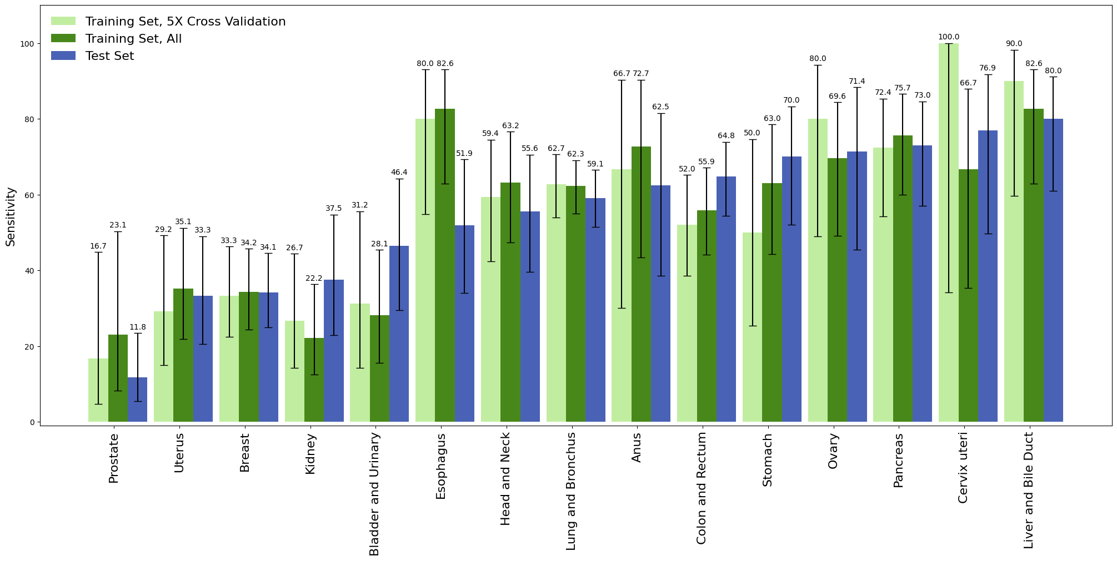
MP, methylation-protein.

Supplementary Figure 4. Evaluation of collection site impact on MP classifier test set performance.
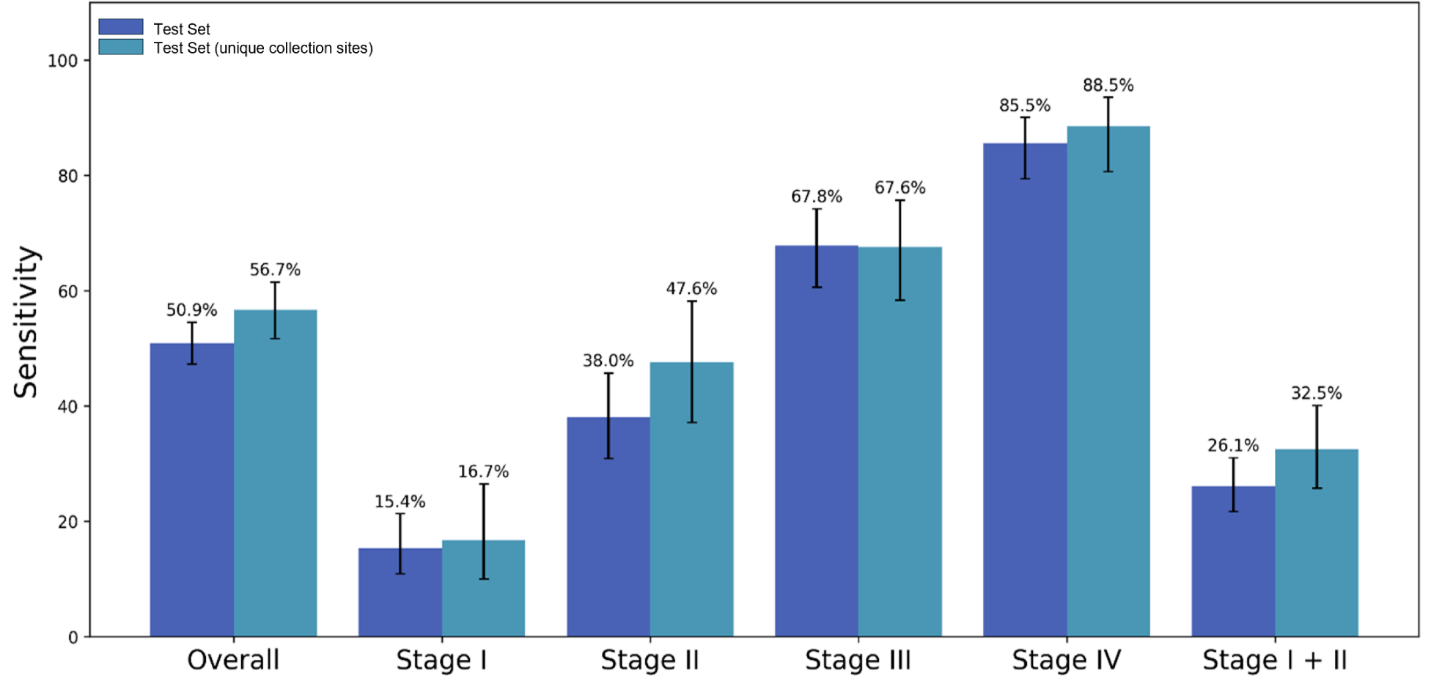

MP, methylation-protein.
